# Human genetic variation associates with infection by derived Ugandan M. tuberculosis lineage

**DOI:** 10.1101/2025.10.31.25339263

**Authors:** Catherine M. Stein, Penelope Benchek, Lentlamatse Mantshoyane, Timothy Ciesielski, Michael L. McHenry, Himiede Wilson-Sesay, Moses Joloba, Eddie Wampande, Kimberly A. Dill Mc-Farland, Allison W. Roberts, Ben Polacco, Max Bennett, Nevan Krogan, W. Henry Boom, Jeffery S. Cox, Harriet Mayanja-Kizza, Thomas R. Hawn, Scott M. Williams

**Author notes:** Address correspondence to: Dr. Catherine Stein. Author approval: All authors have seen and approve of the manuscript.

## Abstract

Several studies examined host and pathogen genetic influences on tuberculosis (TB) susceptibility separately, but relatively few studied their combined effects. However, host-pathogen interactions or co-evolution may explain the inability to replicate many reported human genetic effects across global populations and provide additional insight into TB risk. In this study, we address such possible interactions by focusing on the outcome of infection with L4-Uganda *M. tuberculosis* sub-lineage and human genetic variants as independent variables. This is possible because the L4-Uganda sub-lineage is both restricted to Uganda and nearby locations and is recent there, compared to other more ancestral L4 lineages. Our study consisted of 276 culture-confirmed adult TB cases from a long-standing household contact study. Multiple loci with results suggestive of association (p<10^-5^) also demonstrated convergent relevant evidence for strain specific infection via: evidence of gene expression in relevant cells and lung tissue, signatures of natural selection, eQTL expression, and CRISPR screens for immunity-related genes. We also replicated previously published host-pathogen interaction effects, demonstrating that effects seen for other sub-lineages were also present for L4-Uganda. These results provide evidence for host-pathogen co-evolution in TB and indicate these interactions involve genes highly relevant to the host immune response to *Mycobacterium* infection.

## INTRODUCTION

Tuberculosis (TB) remains a major public health problem globally and is the leading infectious disease killer globally ^1^. Caused by *Mycobacterium tuberculosis* (Mtb), TB disease ultimately results from a host-pathogen interaction, where susceptibility to disease is influenced by both host and microbial genetic factors ^2–5^. The human genetic variants associated with TB often vary by global population. Not only do human genetic associations vary by geography, but Mtb genotypes do as well, and can be organized into 8 major lineages that have distinct geographical distributions and timelines of exposure to human populations ^6,7^. Within these 8 major lineages, there are also recently diverged sub-lineages. As ancient lineages are less likely to be virulent than the modern ones, this diversity of lineages and their historical coexistence with humans has led to the hypothesis that disrupted coevolution between the host and Mtb complex genes increases virulence ^8–13^. Consistent with this hypothesis, several candidate gene and genome-wide studies have been conducted ^4,14–23^, demonstrating host-pathogen interaction effects at the human DNA – Mtb lineage level. However, most of these studies have been done in East Asia and the presence of such interaction effects in Africa are not as thoroughly explored. The role of host-pathogen interactions in Africa is critical as it is thought TB originated on the continent^24^, thereby providing a unique opportunity to assess how interactions between the two species can affect disease risk in light of ancient versus recently diverged, sub-lineages.

In our previous work ^25,26^, we posited that the relatively recent introduction into the Ugandan population of the L4-Uganda sublineage could result in co-evolution with host genotypes that affected disease severity. This is a different question than asking whether human genetic variation affects susceptibility to disease caused by a specific Mtb lineage, which is the approach taken by the other above cited studies. Given that the L4-Uganda sub-lineage is recently diverged from the ancestral L4 lineage and is unique to Uganda and surrounding countries, it is possible to identify novel human genetic variants that convey TB risk caused by this newer L4-Uganda lineage compared to older lineages that co-existed with humans in Uganda for much longer. Thus in this work, in contrast to our prior work on severity, we sought to identify human genetic factors significantly associated with pulmonary TB due to infection by the L4-Uganda sub-lineage. In Kampala, Uganda, both L4-Uganda and L4-NonUganda are common, providing a unique natural experiment in which to test the hypothesis that human genetic variants associate with strain specific disease.

## METHODS

### Subject ascertainment and characterization

As described previously^26^, this study includes TB cases ascertained through a household contact study in Kampala, Uganda from 2002-2012. Culture-confirmed adult TB cases with human genome-wide association (GWAS) data and Mtb lineage data, as described previously ^27^, were included in these analyses. Subjects were grouped into two cohorts, based on which human genotyping chips were accessible at the time of genotyping. We refer to them as Cohort 1 and Cohort 2 (N=149 and N=127, respectively). The study protocol was approved by the National HIV/AIDS Research Committee of Makerere University and the institutional review board at University Hospitals Cleveland Medical Center. Final clearance was given by the Uganda National Council for Science and Technology. All participants provided written informed consent. Additional details about the original study protocol are described elsewhere^28^. The two cohorts differed in percentage of HIV positive individuals (Table 1); therefore, HIV status was used as a covariate in all analyses.

**Table 1.** Sample characteristics.

|  | **Cohort 1** | **Cohort 2** | **Total** | **P** |
| --- | --- | --- | --- | --- |
|  | **N=149** | **N=127** |  |  |
| **Mean Age (SD)** | 28.7 (8.1) | 28.7(9.8) | 28.7 (9.0) | 0.99 |
| **# Male (%)** | 81 (54.4) | 73 (57.5) | 154 (55.8) | 0.69 |
| **# HIV+ (%)** | 31 (24.4) | 15 (10.1) | 46 (16.7) | 0.0025 |
| **Mean TBscore (SD)** | 6.2 (2.1) | 5.4 (2.2) | 5.8 (2.2) | 0.0032 |
| **# L4.6/Ugandan (%)** | 93 (62.4) | 75 (59.1) | 168 (60.9) | 0.49 |
| **# L4 (%)** | 17 (11.4) | 21 (16.5) | 38 (13.7) |  |
| **# L3 (%)** | 37 (24.8) | 31 (24.4) | 68 (24.6) |  |

### Human genotyping and Lineage typing

Genotyping methods have been described in detail elsewhere ^26,29^. Briefly, Cohort 1 was genotyped on the Illumina Infinium MegaEX chip, and Cohort 2 used the Illumina HumanOmni5 microarray. Both cohorts were separately imputed to the TOPMED-r2 reference panel. Prior to imputation, only SNPs that had a call rate greater than 0.98, minor allele frequency (MAF) > 0.05, and did not show deviation from Hardy-Weinberg equilibrium (p<10^−6^) were retained. The total number of well-imputed (R^2^ > 0.8) SNPs that overlapped between the two cohorts was 18,468,458 (7,925,870 at MAF > 5%). Principal components were computed using PCAiR^30^.

Mtb was isolated from sputum of each of the subjects, and lineages were classified according to lineage-identifying SNPs using real-time PCR and validated with long sequence polymorphism (LSP) PCR as described previously ^27^. Lineage was determined using three SNPs that accurately distinguish all lineages found in Uganda, L4.6 Uganda (aka L4-Uganda), L3, and L4-NonUganda lineages ^6,7^. L4-Uganda is a sub-lineage that recently diverged from the parent L4 lineage that is only found in Uganda and countries immediately surrounding it ^6,7,9,31^.

### GWAS analysis

In our primary analysis, we assessed as outcomes disease due to L4-Uganda vs. all other L4 lineages; this analysis contrasts the recently evolved sub-lineage with the “older” sub-lineages of the same phylogenetic family ^6,9^ GWAS analyses were performed in the two separate cohorts on well-imputed (R^2^ > 0.8) SNPs with a within cohort minor allele frequency (MAF) > 5% and a within cohort minor allele count (MAC) ≥ 15 using the penalized quasi-likelihood approximation to the generalized linear mixed model in the R, Genesis package. Covariate adjustment included age, sex, HIV status and the first two principal components. A fixed effects meta-analysis was run over the two GWAS utilizing the metafor package in R.

As a sensitivity analysis, we contrasted L4-Uganda with other L4 lineages and the L3 lineage combined. Unlike our primary analysis, this analysis does not strictly capture recently evolved vs older lineages, as this also includes L3, which is present in other global populations and diverged from L4 much longer ago^6,32^.

### Identification of regions with signatures of selection

We examined SNPs with association p<10^-5^ for signatures of natural selection using the extended haplotype homozygosity (EHH) approach ^33^. Under this model, variants under recent positive selection will demonstrate a large region of surrounding haplotype homozygosity. We leveraged a newer measure based on EHH that accounts for local recombination hotspots: nSL scores (number of segregating sites by length)^34^. We examined whole genome sequence data available in the 1000 Genomes database (phased 30x coverage build 38 data)^35^, using Selscan 2.0 to obtain 26 nSL scores for each SNP (one nSL score per 1000 Genome population). Then for each population, we normalized the nSL scores in 100 bins of similar frequency to adjust for the impact of allele frequency on mean EHH. Finally, we evaluated if any of these normalized |nSL| scores were in the top 1% of scores for the population they were measured in. SNPs with a top1% |nSL| score were considered to be under recent positive selection in that population.

### Bioinformatic annotation

We further investigated our most significant findings using a number of bioinformatic annotation tools. We prioritized SNPs that were significant at p<10^-6^ and any other suggestive SNPs that showed signatures of selection as identified above. First, we used HumanBase (formerly known as GIANT^36^) to identify tissues and cell types where proximal genes were expressed and how these genes were connected in gene expression networks. Second, we used FAVOR^37^ to identify whether associated SNPs had epigenetic features, transcription factor binding sites, showed conservation across species according to GERP scores, and potential gene function implications through CADD scores. Third, we queried the most significantly associated genes from each region in the GWAS catalog for evidence of pleiotropy^38^.

### Query of eQTLs and CRISPR screens

In a previous study ^39^ we identified SNPs that were associated with differential gene expression in response to macrophage stimulation with *M. tuberculosis in vitro*. We queried the regions associated with Mtb lineage at the p<10^-5^ level for evidence of such eQTLs (Table 2). In addition, a recent genome-wide CRISPR knockout screen in cells ^40^ was conducted independently, to identify macrophage genes that influence the induction of TNF and iNOS upon infection with Mtb. We also queried genes attaining p<10^-6^, a gene identified with a selection signature, and the eQTLs (Table 2) in this dataset.

**Table 2.** Evidence for loci with p < 10^-5^ and at least one other point of supportive evidence.

| locus | Position<br>(build38) | top<br>gene(s) | $p < 10^{-6}$ | signature<br>of<br>selection | expression<br>in relevant<br>tissues | pathway<br>association | eQTL<br>($p < 0.05$) | CRISPR<br>screen<br>p-value | GERP<br>score | relevant<br>associated traits<br>in GWAS catalog<br>(for top genes) |
| --- | --- | --- | --- | --- | --- | --- | --- | --- | --- | --- |
| chr 1 | 156760371<br>156765602<br>156776463<br>156776816 | *PRCC*,<br>*HDGF* | yes |  | yes | in network<br>with<br>SLC8B1 | *RRNAD1* |  | 4.27 | neutrophil<br>count, leukocyte<br>quantity |
| chr2 | 148613271 | (none) | yes |  | N/A |  |  |  |  | N/A |
| chr<br>12 | 91916625<br>113783216<br>113783825 | *SLC8B1* |  | yes |  | in network<br>with chr 1<br>genes |  |  |  |  |
| chr<br>16 | 840155<br>840260<br>840308<br>840327 | *LMF1* | yes |  |  |  | *NARFL*/<br>*CIAO3* | 0.026<br>(*NARFL*<br>) | 6.3 | diabetes |

### Examination of loci reported in prior studies

We first examined association with previously-identified TB candidate genes, as summarized in our previous review^2^. Second, we examined loci that have been implicated in host-pathogen interaction analyses in other studies ^14–23^. Because these are hypothesis-driven queries, we did not apply any multiple testing correction.

## RESULTS

The sample of TB cases was split into two independent cohorts, as described previously^26^; the first cohort had 149 TB cases, and the second had 127 cases (Table 1). The cohorts differed in their prevalence of HIV-infection and TB severity based on the Bandim TBscore, so HIV status was retained as a covariate in the GWAS. We did not adjust for disease severity as our previous work in this same dataset has shown there is no association between lineage and TBscore ^25,26^.

### GWAS Results

In the meta-analysis, ten SNPs associated with disease due to L4-Uganda infection at p < 10^-6^ and an additional 81 SNPs at p < 10^-5^ (Figure 1, Supplemental Table 1). Among the regions that attained significance at the p<10^-6^ threshold, there were four SNPs on chromosome 1 (∼156 Mb) (Figure 2A) with an additional six SNPs in the region significant at p<10^-5^. On chromosome 2 (Figure 2B), there was one SNP significant at p<10^-6^ (∼149 Mb) with one flanking SNP with p<10^-5^ in this gene poor region. In addition, there were four SNPs on chromosome 16 (∼890 kb) with p<10^-6^ with an additional 11 flanking SNPs with p<10^-5^ (Figure 2C). Furthermore, one SNP on chromosome 9 had a SNP meeting the p < 10^-6^ threshold but there were no other associated SNPs in that region at even the suggestive threshold (p<10^-5^), so this region was not considered further. SNPs on chromosome 12 barely missed the p<10^-6^ threshold, with most significant p=1.1×10^-6^ and another with p=1.2×10^-6^ (Figure 2D).

**Figure 1.**
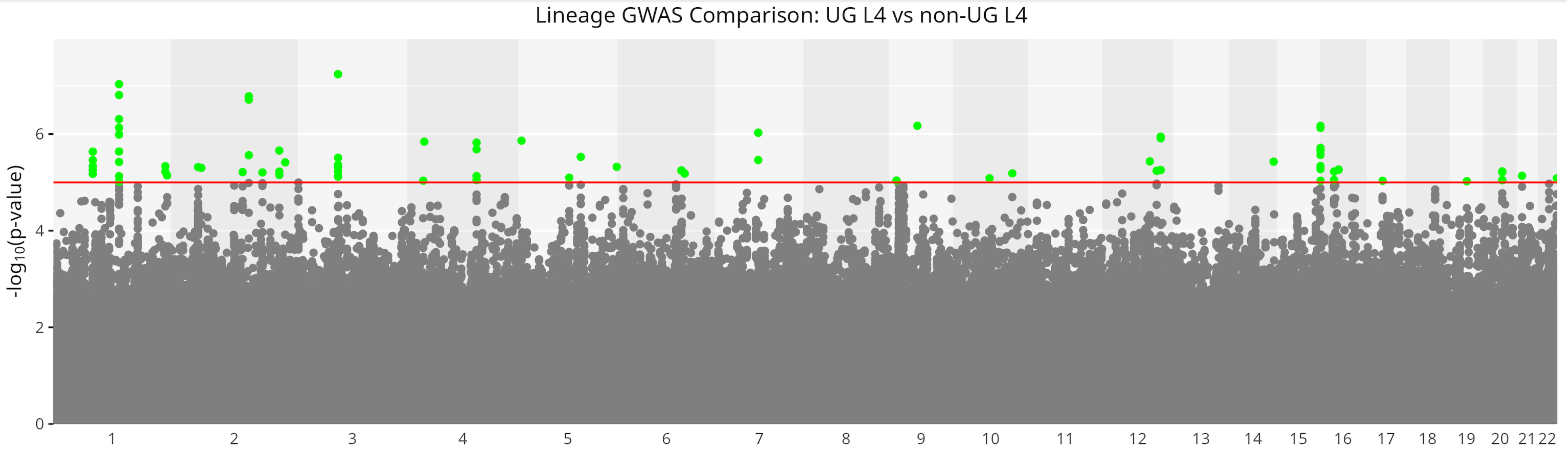
Manhattan plot

**Figure 2.**
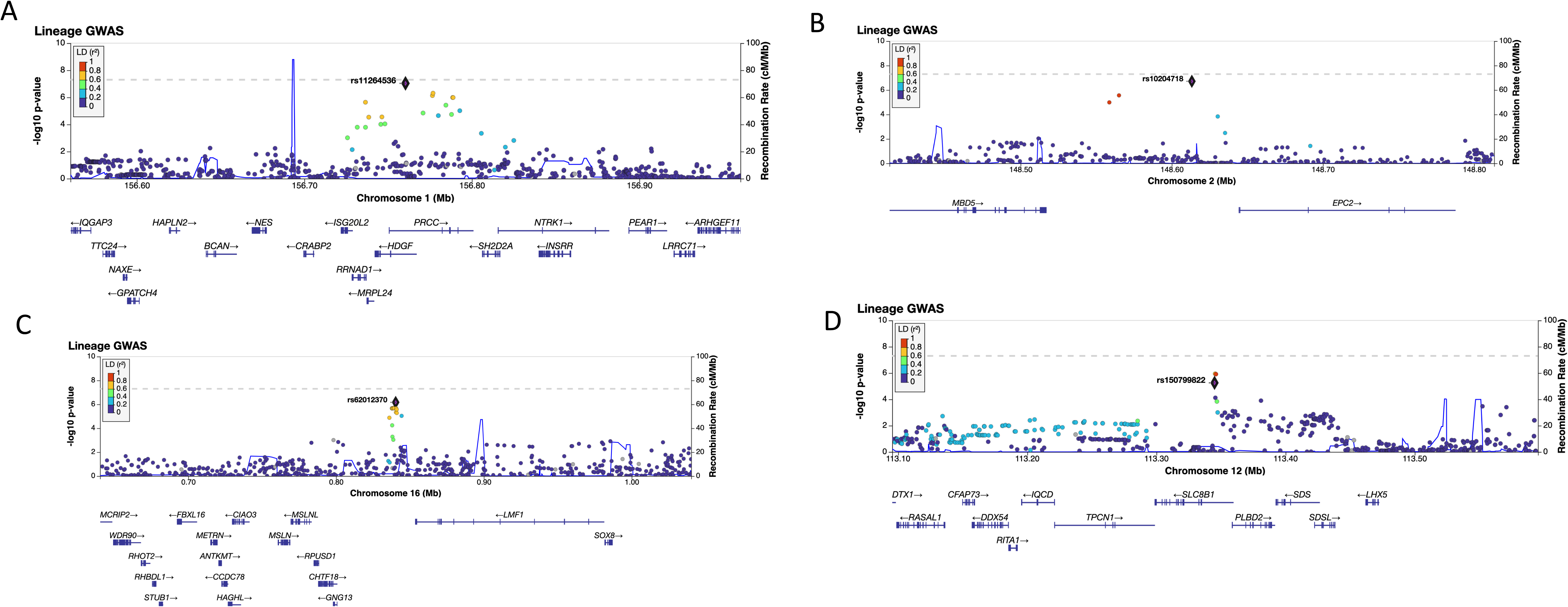
LocusZoom plots for most significant loci: A) Chromosome 1; B) Chromosome 2; C) Chromosome 16; D) Chromosome 12

We examined all of the SNPs yielding association p<10^-5^ for selection signatures in each of the 1000 Genome populations independently. Four of the 81 SNPs demonstrated recent evidence of positive selection in at least one of the 26 populations. All these SNPS were located within the same region on chromosome 12 (within an intron of *SLC8B1*). This region was only under selection in African populations (6 of 7 African populations; Supplemental Table 2). The one African population that showed no evidence of selection in this region was the Mandinka of the Gambian Western Division.

### Bioinformatic annotation

We focused our next analyses using bioinformatic annotation and eQTL queries on the three loci attaining significance at the p<10^-6^ threshold plus the one locus showing both evidence of selection that had a suggestive association (Table 2). The most significant SNP associations on chromosome 1 mapped to two genes with overlapping coding regions, *PRCC* and *HDGF* (Figure 2A). Gene expression profiles in HumanBase revealed that these genes are highly expressed in lung, leukocytes, and lymphocytes, and are expressed at a lower level in monocytes, and neutrophils, (Figure 3A). A network analysis (Figure 3B) revealed that these genes are co-expressed with several genes key to the TB immune response, including *IL12B*, *IL12A*, and *TNF*, along with *SLC8B1*, the latter of which was the gene shown to have signatures of selective pressure in African populations (Supplemental Table 2, Figure 2D). The GWAS catalog revealed that *PRCC* and *HDGF* are associated with neutrophil count and leukocyte quantity, and the GERP score indicates potential functional importance of this SNP. Lastly, our eQTL query revealed a significant eQTL in this region for *RRNAD1* (aka *METTL25B*) (Supplemental Table 3). This gene appears in our network diagram (Figure 3B). We further examined gene expression profiles in HumanBase, and found this gene was also highly expressed in lung and leukocytes (Figure 3).

**Figure 3.**
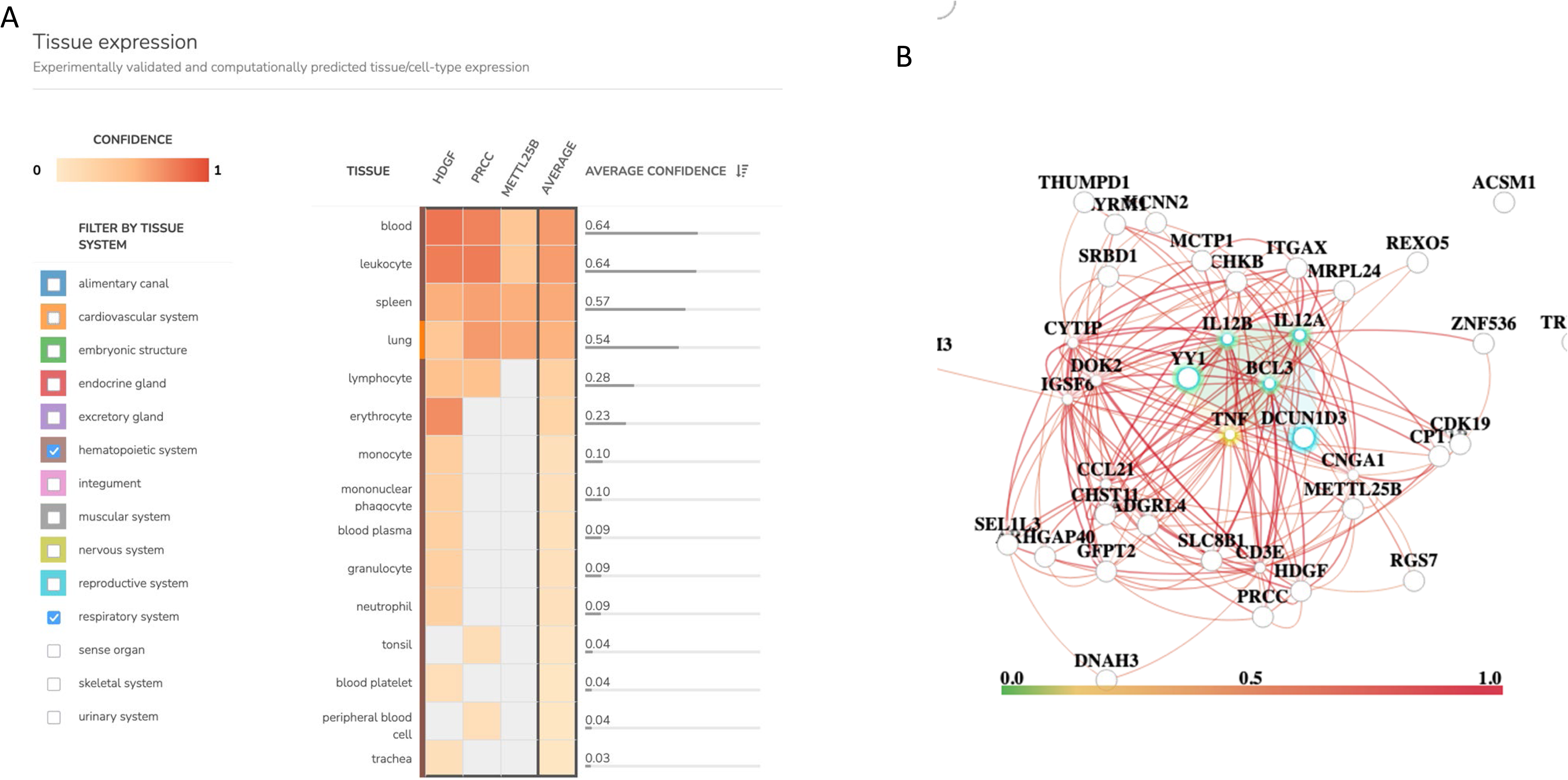
Gene expression profiling from HumanBase for genes on chromosome 1: a) tissue and cell type expression, and b) network diagram depicting co-expression with other genes

Another region, on chromosome 16, showed that the associated SNPs in closest proximity to *LMF1* (Figure 2C); the most closely related phenotype in the GWAS catalog for this gene was diabetes (Table 2). While this gene is not highly expressed in any tissues or cells of interest (data not shown), our eQTL query revealed an association with *NARFL* (aka *CIAO3*) (Supplemental Table 3). The associated gene also had a high GERP score (6.3), indicating functional importance. In addition, the CRSPR macrophage screen showed a tentative association with *NARFL* (p=0.0266, data not shown) ^40^; two of the four guides that targeted *NARFL* were enriched in the TNF+iNOS-population. These results did not attain significance after multiple testing correction.

### Query of previously studied TB candidate genes and host-pathogen interaction effects

First, we examined whether any candidate genes previously associated with TB susceptibility ^2^ were associated with infection by L4-Uganda lineage (Supplemental Table 4). While some of these results fell within type I error rate (proportion of SNPs with p<0.05 was ∼5%), there were some candidate genes with well more than 5% of SNPs that reached a p threshold of 0.05.

*SLC11A1*, aka *NRAMP1*, was associated with infection by L4-Uganda (most significant SNP p=0.0055; 19% of SNPs reached p < 0.05); interestingly, this gene was previously indicated in an interaction effect with L4-Uganda in its association with disease severity as measured by the Bandim TBscore ^25^ along with susceptibility to infection by Beijing lineage ^21^. Similarly, SNPs in *TLR2* were associated with infection with L4-Uganda (p=0.0043); this gene has previously been associated with infection with Beijing lineage in Vietnam^16^. Another highly significant association was the vitamin D receptor gene (most significant p=0.007).

Second, we queried genes and regions that were previously implicated in similar host-Mtb lineage interaction studies (Supp Table 5). While many results approached significance at p<0.10, a few loci were notably more significant. Many of these have been previously associated with infection with the Beijing lineage, including *CD53* (p=0.000003), *HLA-B* (p=0.0006), as well as the L2.2 lineage that is likely related to Beijing, including FSTL5 (p=0.0035), *CSGAL/NCAT1* (p=0.0061), along with one gene associated with an L1 sub-lineage (*RIMS3*, p=0.0077).

### Sensitivity analysis including L3 lineage

Lastly, we conducted a sensitivity analysis to evaluate the impact of including L3 lineage within the referent group of the outcome variable. There were too few subjects with L3 lineage for this to be considered a separate contrast (Table 1). Thus, we examined the correlation of p-values from our primary analysis, with those found when L4-Uganda was contrasted with both L4-nonUganda and L3 (Supplemental Figure 1). As shown in the correlation plot, the p-values at the most significant loci detected by the L4-Uganda vs L4-NonUganda GWAS had p-values in the L4-Uganda vs L4-NonUganda plus L3 GWAS that were barely significant at p<0.001, orders of magnitude less significant that the initial finding for L4-Uganda versus L4. In other words, these loci would not have been detected at the “suggestive” p<10^-5^ threshold if the L3 lineage was included in the reference category. This analysis reveals that inclusion of the historically-diverged primary lineage of L3 decreases power, and that any co-evolutionary effects may have been missed if the contrast was not between the recently evolved lineage and its parent phylogenetic branch.

## DISCUSSION

Our objective in this analysis was to further examine the paradigm of host-pathogen interaction in TB susceptibility, by examining the human genome for variants associated with disease due to the recently evolved L4-Uganda lineage. Under the co-evolutionary model that we have previously proposed ^25,26^, SNPs associated with L4-Uganda versus the older L4-NonUganda would provide evidence of disruptive co-evolution. Our genome-wide analysis revealed five loci attaining suggestive signals. Two of these are in loci that are eQTLs for genes exhibiting differential expression in response to *M. tuberculosis* stimulation *in vitro*, and a third shows signatures of recent natural selection. In addition, we replicated previously reported genes involved in host-pathogen interaction effects as well as TB susceptibility genes. Together, these results demonstrate that the effect of immune response genes on TB susceptibility depends on the context of the infecting Mtb lineage. A unique strength of this work is the availability of data on eQTLs that are specific to the *in vitro* response to *M. tuberculosis* stimulation, as well as the availability of results from a genome-wide CRISPR screen of macrophage response to Mtb, to evaluate the biologic plausibility of our findings. Another unique strength of this work is the ability to contrast a recently diverged lineage from its ancestral lineage, pointing to potential co-evolutionary effects. Overall, our results provide further support for the concept that the effect of host genes on TB susceptibility may depend on the genetic background of the pathogen and the historical concordance between human alleles and lineage. This work demonstrates that these interaction effects are most prominent in immune response genes.

Results for chromosome 1 showed several associated SNPs – two of these map to genes *PRCC* and *HDGF*, and these SNPs are eQTLs for the nearby gene *RRNAD1*. Interestingly, these genes are all expressed in lung and immune cells and are also connected in a co-expression network in dendritic cells along with other genes that are part of the immune response to TB. Both *PRCC* and *HDGF* have been associated with neutrophil count in prior GWAS studies; neutrophils and dendritic cells play key roles in the immune response to TB^41,42^. Thus, our results support the hypothesis that genes in this region have a role in disease susceptibility most likely acting through host-pathogen interactions. There is extensive linkage disequilibrium in this region, and network analysis indicates these genes are co-expressed, so it is difficult to discern which gene(s) are causal, but as most significant the SNP is an eQTL for *RRNAD1,* it is a strong candidate.

Chromosome 16 also includes some promising candidate genes. *NARFL* was weakly associated in the CRISPR screen, indicating a role in regulating early innate immune responses during the initial interaction of macrophages and Mtb. These results, though tentative, indicate that decreased *NARFL* results in more TNF production, so *NARFL* may inhibit TNF. *NARFL* (aka *CIAO3*) codes for cytosolic iron-sulfur assembly component 3, and recent work showed that iron-sulfur cofactors are essential for intracellular adaption of Mtb^43^. The role of bioelements such as iron and sulfur is currently an active area of TB research, thus making this gene another promising candidate for further investigation. On the other hand, the connection between *LMF1* and TB susceptibility is less clear.

We also examined genes that have been previously associated with TB susceptibility as well as those previously implicated in host-pathogen interaction effects on disease risk and severity.

Several of these genes appeared to be overrepresented in terms of the number of SNPs that associated with TB in our data, providing evidence for replication. Most notably, we found several immune genes that were associated with L4-Uganda infection, including *NRAMP1 (SLC11A1), TLR2, HLA-B*, and *CD53*. As we noted in our prior work ^25^, *NRAMP1* is the most studied TB candidate gene, but association results have been inconsistent across world populations. Our current findings, along with others ^21^, indicate that the effect of *NRAMP1* is dependent on pathogen lineage, which varies globally. While there has been a great emphasis on *NRAMP1* in prior literature, this current work reveals that other immune genes are also subject to host-pathogen effects. The idea that immunological responses to TB may be pathogen-dependent^44,45^ is critical for understanding how to best design future vaccine and other therapeutic targets.

A limitation of this study is the sample size. This restricted our ability to examine effects of the entire L4 lineage vs L3 and may have resulted in reduced power to replicate other previously-reported genetic effects. While our work focused on one recently diverged L4 sub-lineage, it is likely that similar effects might be seen for other Mtb sub-lineages elsewhere. In addition, our bacterial genetic data are limited to the lineage level, so these results cannot be translated to effects due to specific genetic variants in Mtb.

In conclusion, this work provides further support for host genetic effects that are significant primarily in the context of specific Mtb genetic backgrounds. Both our novel findings, and replication of previously published findings, demonstrate that these host genes tend to have roles in immunity or other relevance to Mtb infection and progression to disease. Furthermore, the results indicate that replication of host genetic associations across global populations may not be expected when such co-evolutionary effects exist. Ancestry explains genetic differences in immunity-related genes ^46^, and genetic variation in immunity-related genes has been shaped by natural selection. Finally, genetic epidemiological studies need to incorporate cross species genetic effects to inform functional investigation in the pursuit of better treatment outcomes for TB.

## ACKNOWLEDGEMENTS

We want to acknowledge the contributions made by senior physicians, medical officers, health visitors, laboratory and data personnel: Dr. Lorna Nshuti, Dr. Roy Mugerwa, Dr. Alphonse Okwera, Dr. Deo Mulindwa, Dr. Mary Nsereko, Denise Johnson, Dr. Allan Chiunda, Hussein Kisingo, Mark Breda, Dennis Dobbs, Mary Rutaro, Albert Muganda, Richard Bamuhimbisa, Yusuf Mulumba, Deborah Nsamba, Barbara Kyeyune, Faith Kintu, Gladys Mpalanyi, Janet Mukose, Grace Tumusiime, Pierre Peters, Annet Kawuma, Saidah Menya, Joan Nassuna, Keith Chervenak, Karen Morgan, Alfred Etwom, Micheal Angel Mugerwa, Emily Hellwig, and Lisa Kucharski. We would like to acknowledge Dr. Francis Adatu Engwau, former Head of the Uganda National Tuberculosis and Leprosy Program, for supporting this project. We would like to acknowledge the medical officers, nurses and counselors at the National Tuberculosis Treatment Centre, Mulago Hospital, the Ugandan National Tuberculosis and Leprosy Program and the Uganda Tuberculosis Investigation Bacteriological Unit, Wandegeya, for their contributions to this study.

## Data availability statement

Due to Ugandan IRB restrictions, these data cannot be deposited publicly. Requests for data can be made to the chair of the data access committee, Assoc Prof Erisa Sabakaki Mwaka.

## Funding Statement

Funding for this work was provided by U19AI162583, N01 AI95383, R01HL096811, T32HL007567, and R01AI124348.

Supplemental Table 1 – results with p < 10^-5^

Supplemental Table 2 – results from selection signature analysis

Supplemental Table 3 – eQTL query

Supplemental Table 4 – Query of TB susceptibility candidate genes from the literature

Supplemental Table 5 – Query of genes previously identified in host-pathogen interaction studies of TB

Supplemental Figure 1 – correlation of p-values for L4-Uganda / L4-NonUganda GWAS vs. L4-Uganda / all other lineages

## References

1. World Health Organization. Global tuberculosis report 2024. (https://www.who.int/teams/global-programme-on-tuberculosis-and-lung-health/tb-reports/global-tuberculosis-report-2024, 2004).

2. Stein, C.M. et al. Genomics of human pulmonary tuberculosis: from genes to pathways. Curr Genet Med Rep (2017).

3. Schurz, H. et al. Multi-ancestry meta-analysis of host genetic susceptibility to tuberculosis identifies shared genetic architecture. medRxiv, 2022.08.26.22279009 (2022).

4. Ndong Sima, C.A.A., et al. The immunogenetics of tuberculosis (TB) susceptibility. Immunogenetics (2022).

5. McHenry, M.L., Williams, S.M. & Stein, C.M. Genetics and evolution of tuberculosis pathogenesis: New perspectives and approaches. Infect Genet Evol 81, 104204 (2020).

6. Stucki, D. et al. Mycobacterium tuberculosis lineage 4 comprises globally distributed and geographically restricted sublineages. Nat Genet 48, 1535–1543 (2016).

7. Ngabonziza, J.C.S. et al. A sister lineage of the Mycobacterium tuberculosis complex discovered in the African Great Lakes region. Nat Commun 11, 2917 (2020).

8. Gagneux, S. & Small, P.M. Global phylogeography of Mycobacterium tuberculosis and implications for tuberculosis product development. Lancet Infect Dis 7, 328–37 (2007).

9. Gagneux, S. Ecology and evolution of Mycobacterium tuberculosis. Nat Rev Microbiol 16, 202–213 (2018).

10. Coscolla, M. & Gagneux, S. Consequences of genomic diversity in Mycobacterium tuberculosis. Seminars in immunology 26, 431–444 (2014).

11. Comas, I. et al. Out-of-Africa migration and Neolithic coexpansion of Mycobacterium tuberculosis with modern humans. Nat Genet 45, 1176–82 (2013).

12. Gagneux, S. Host-pathogen coevolution in human tuberculosis. Philosophical transactions of the Royal Society of London. Series B, Biological sciences 367, 850–859 (2012).

13. Kodaman, N., Sobota, R.S., Mera, R., Schneider, B.G. & Williams, S.M. Disrupted human-pathogen co-evolution: a model for disease. Front Genet 5, 290 (2014).

14. Luo, Y. et al. Paired analysis of host and pathogen genomes identifies determinants of human tuberculosis. Nat Commun 15, 10393 (2024).

15. Salie, M. et al. Associations between human leukocyte antigen class I variants and the Mycobacterium tuberculosis subtypes causing disease. J Infect Dis 209, 216–23 (2014).

16. Caws, M. et al. The influence of host and bacterial genotype on the development of disseminated disease with Mycobacterium tuberculosis. PLoS Pathog 4, e1000034 (2008).

17. Thuong, N.T. et al. MARCO variants are associated with phagocytosis, pulmonary tuberculosis susceptibility and Beijing lineage. Genes Immun 17, 419–425 (2016).

18. Intemann, C.D. et al. Autophagy gene variant IRGM −261T contributes to protection from tuberculosis caused by Mycobacterium tuberculosis but not by M. africanum strains. PLoS Pathog 5, e1000577 (2009).

19. Thye, T. et al. Variant G57E of mannose binding lectin associated with protection against tuberculosis caused by Mycobacterium africanum but not by M. tuberculosis. PLoS One 6, e20908 (2011).

20. Songane, M. et al. Polymorphisms in autophagy genes and susceptibility to tuberculosis. PLoS One 7, e41618 (2012).

21. van Crevel, R. et al. Infection with Mycobacterium tuberculosis Beijing genotype strains is associated with polymorphisms in SLC11A1/NRAMP1 in Indonesian patients with tuberculosis. J Infect Dis 200, 1671–4 (2009).

22. Omae, Y. et al. Pathogen lineage-based genome-wide association study identified CD53 as susceptible locus in tuberculosis. J Hum Genet 62, 1015–1022 (2017).

23. Toyo-Oka, L. et al. Strain-based HLA association analysis identified HLA-DRB1*09:01 associated with modern strain tuberculosis. Hla 90, 149–156 (2017).

24. Barberis, I., Bragazzi, N.L., Galluzzo, L. & Martini, M. The history of tuberculosis: from the first historical records to the isolation of Koch’s bacillus. J Prev Med Hyg 58, E9–e12 (2017).

25. McHenry, M.L. et al. Interaction between host genes and Mycobacterium tuberculosis lineage can affect tuberculosis severity: Evidence for coevolution? PLoS Genet 16, e1008728 (2020).

26. McHenry, M.L. et al. Interaction between M. tuberculosis Lineage and Human Genetic Variants Reveals Novel Pathway Associations with Severity of TB. Pathogens 10(2021).

27. Wampande, E.M. et al. A single-nucleotide-polymorphism real-time PCR assay for genotyping of Mycobacterium tuberculosis complex in peri-urban Kampala. BMC Infect Dis 15, 396 (2015).

28. Stein, C.M. et al. Resistance and susceptibility to Mycobacterium tuberculosis infection and disease in tuberculosis households in Kampala, Uganda. Am J Epidemiol 187, 1477–1489 (2018).

29. McHenry, M.L. et al. Tuberculosis severity associates with variants and eQTLs related to vascular biology and infection-induced inflammation. medRxiv, 2022.08.23.22279140 (2022).

30. Conomos, M.P., Miller, M.B. & Thornton, T.A. Robust inference of population structure for ancestry prediction and correction of stratification in the presence of relatedness. Genet Epidemiol 39, 276–93 (2015).

31. Gagneux, S. Host-pathogen coevolution in human tuberculosis. Philos Trans R Soc Lond B Biol Sci 367, 850–9 (2012).

32. Brites, D. & Gagneux, S. The Nature and Evolution of Genomic Diversity in the Mycobacterium tuberculosis Complex. Adv Exp Med Biol 1019, 1–26 (2017).

33. Sabeti, P.C. et al. Detecting recent positive selection in the human genome from haplotype structure. Nature 419, 832–837 (2002).

34. Ferrer-Admetlla, A., Liang, M., Korneliussen, T. & Nielsen, R. On detecting incomplete soft or hard selective sweeps using haplotype structure. Mol Biol Evol 31, 1275–91 (2014).

35. Byrska-Bishop, M. et al. High-coverage whole-genome sequencing of the expanded 1000 Genomes Project cohort including 602 trios. Cell 185, 3426–3440.e19 (2022).

36. Greene, C.S. et al. Understanding multicellular function and disease with human tissue-specific networks. Nat Genet 47, 569–76 (2015).

37. Zhou, H. et al. FAVOR: functional annotation of variants online resource and annotator for variation across the human genome. Nucleic Acids Res 51, D1300–d1311 (2023).

38. Cerezo, M. et al. The NHGRI-EBI GWAS Catalog: standards for reusability, sustainability and diversity. Nucleic Acids Res 53, D998–d1005 (2025).

39. Hong, H. et al. Mycobacterium tuberculosis-dependent monocyte expression quantitative trait loci, cytokine production, and TB pathogenesis. Front Immunol 15, 1359178 (2024).

40. Roberts, A.W.D.C., L.N.; Garelis, N.E.; Cox, J.S. Genome-wide screen in Mycobacterium tuberculosis infected macrophages reveals innate regulation of antibacterial mediators by IRF2. https://www.biorxiv.org/content/10.1101/2025.09.26.678671v1 (2025).

41. Rahman, F. Characterizing the immune response to Mycobacterium tuberculosis: a comprehensive narrative review and implications in disease relapse. Front Immunol 15, 1437901 (2024).

42. Kroon, E.E. et al. Neutrophils: Innate Effectors of TB Resistance? Front Immunol 9, 2637 (2018).

43. Pandey, M., Talwar, S., Bose, S. & Pandey, A.K. Iron homeostasis in Mycobacterium tuberculosis is essential for persistence. Sci Rep 8, 17359 (2018).

44. Wang, C. et al. Innate immune response to Mycobacterium tuberculosis Beijing and other genotypes. PLoS One 5, e13594 (2010).

45. Tientcheu, L.D. et al. Immunological consequences of strain variation within the Mycobacterium tuberculosis complex. Eur J Immunol 47, 432–445 (2017).

46. Sanz, J., Randolph, H.E. & Barreiro, L.B. Genetic and evolutionary determinants of human population variation in immune responses. Curr Opin Genet Dev 53, 28–35 (2018).

