## Supplemental Table and Figure for "Human genetic variation associates with infection by derived Ugandan M. tuberculosis lineage"

SUPPLEMENTAL TABLES

**Supplemental Table 1. SNPs associated with L4-Uganda lineage at p<10-5 threshold**

| chr | Position (build 38) | rsID | Ref_allele | Alt_allele | gwasP | beta | se | MAF |
|---|---|---|---|---|---|---|---|---|
| 1 | 79077303 | rs61522925 | C | CT | 5.62E-06 | -0.97469 | 0.214674 | 0.4153 |
| 1 | 79080689 | rs4311840 | A | T | 6.64E-06 | -0.96576 | 0.214381 | 0.4153 |
| 1 | 79081588 | rs113330397 | A | AG | 3.46E-06 | -0.98832 | 0.212934 | 0.4281 |
| 1 | 79087092 | rs10874010 | G | A | 2.31E-06 | -0.99774 | 0.211184 | 0.4281 |
| 1 | 79087986 | rs7533890 | G | A | 4.64E-06 | -0.97971 | 0.21388 | 0.4153 |
| 1 | 156736467 | rs12036794 | T | C | 2.29E-06 | 1.344341 | 0.284471 | 0.1286 |
| 1 | 156760371 | rs11264536 | A | G | 9.25E-08 | 1.448959 | 0.271295 | 0.152 |
| 1 | 156765602 | rs11587467 | A | G | 1.55E-07 | 1.195623 | 0.227912 | 0.2965 |
| 1 | 156776463 | rs34667125 | T | TAG | 7.38E-07 | 1.219986 | 0.24641 | 0.2179 |
| 1 | 156776816 | rs1412310 | A | G | 4.90E-07 | 1.223313 | 0.243187 | 0.2179 |
| 1 | 156783065 | rs10082023 | T | C | 7.42E-06 | 1.056127 | 0.235672 | 0.3404 |
| 1 | 156784486 | rs11264551 | A | G | 3.78E-06 | 1.18531 | 0.256383 | 0.1868 |
| 1 | 156788545 | rs56338469 | C | T | 1.02E-06 | 1.213208 | 0.248227 | 0.2179 |
| 1 | 156788810 | rs111896519 | T | C | 1.03E-06 | 1.213089 | 0.248265 | 0.2179 |
| 1 | 156792798 | rs11589028 | C | A | 9.68E-06 | 1.098494 | 0.248297 | 0.2247 |
| 1 | 241189953 | rs58474067 | A | T | 4.62E-06 | -1.42872 | 0.311851 | 0.1399 |

| | | | | | | | | |
|---|---|---|---|---|---|---|---|---|
| 1 | 241189976 | rs56809646 | G | A | 5.99E-06 | -1.41313 | 0.312175 | 0.1399 |
| 1 | 244250228 | rs56879263 | A | C | 7.18E-06 | -1.53216 | 0.341368 | 0.1505 |
| 2 | 45554214 | rs76565989 | C | T | 4.83E-06 | -1.49419 | 0.326802 | 0.1551 |
| 2 | 50661834 | rs77009079 | T | C | 5.00E-06 | -1.42413 | 0.311986 | 0.1823 |
| 2 | 136453421 | rs578935 | C | T | 6.13E-06 | -1.48583 | 0.328593 | 0.09531 |
| 2 | 148565070 | rs2034268 | A | G | 2.74E-06 | -1.37237 | 0.292672 | 0.1634 |
| 2 | 148613271 | rs10204718 | A | G | 1.93E-07 | -1.52299 | 0.292571 | 0.1672 |
| 2 | 211316552 | rs7607022 | G | A | 5.99E-06 | -1.52759 | 0.337447 | 0.1778 |
| 2 | 211320435 | rs73081744 | A | G | 6.99E-06 | -1.55585 | 0.346205 | 0.1551 |
| 2 | 211321562 | rs151103011 | C | CATT | 2.19E-06 | -1.62669 | 0.343549 | 0.1725 |
| 2 | 221354899 | rs6734155 | A | G | 3.86E-06 | -1.31871 | 0.285495 | 0.2118 |
| 3 | 70726308 | rs10658069 | CAG | C | 5.93E-06 | -1.56746 | 0.34611 | 0.1271 |
| 4 | 25764088 | rs28753900 | A | G | 9.22E-06 | 1.043212 | 0.235244 | 0.3238 |
| 4 | 27903914 | rs6841769 | A | C | 1.44E-06 | -1.10495 | 0.229262 | 0.27 |
| 4 | 118829390 | rs13145450 | C | T | 8.96E-06 | -1.42227 | 0.320267 | 0.1097 |
| 4 | 118836988 | rs67882376 | A | G | 7.40E-06 | -1.41682 | 0.316118 | 0.112 |
| 4 | 118841796 | rs2162400 | G | T | 1.50E-06 | -1.44062 | 0.299421 | 0.09607 |
| 4 | 118845233 | rs34371198 | C | G | 2.07E-06 | -1.44063 | 0.303514 | 0.09531 |
| 5 | 4375909 | rs261129 | T | C | 1.37E-06 | -1.3132 | 0.271904 | 0.2216 |
| 5 | 95056061 | rs13359900 | C | T | 7.96E-06 | -1.75766 | 0.393537 | 0.1301 |
| 5 | 114498966 | rs3891371 | G | A | 2.97E-06 | -1.10703 | 0.236892 | 0.4206 |
| 5 | 180318666 | rs114797869 | T | C | 4.77E-06 | -1.72624 | 0.377354 | 0.0885 |
| 6 | 110596656 | rs114245171 | A | G | 5.67E-06 | -1.40901 | 0.310471 | 0.1036 |
| 6 | 111621048 | rs12526064 | G | T | 5.77E-06 | -1.32601 | 0.292419 | 0.2005 |

| | | | | | | | | |
|---|---|---|---|---|---|---|---|---|
| 6 | 111621101 | rs12524233 | A | G | 5.69E-06 | -1.3259 | 0.292195 | 0.2005 |
| 9 | 9001075 | rs10977434 | T | G | 9.15E-06 | -1.24971 | 0.281703 | 0.1884 |
| 9 | 70979711 | rs11142658 | C | A | 6.73E-07 | -1.26815 | 0.255206 | 0.3275 |
| 10 | 63638031 | rs16918709 | G | C | 8.24E-06 | -1.39373 | 0.312568 | 0.1006 |
| 10 | 107259302 | rs56348641 | T | C | 6.49E-06 | -1.10761 | 0.24561 | 0.2693 |
| 12 | 91522849 | rs75164167 | G | T | 3.68E-06 | 1.400001 | 0.302451 | 0.1248 |
| 12 | 104612404 | rs10861254 | T | C | 5.76E-06 | -1.05373 | 0.232359 | 0.2277 |
| 12 | 113344819 | rs141837440 | A | G | 5.54E-06 | -1.40741 | 0.309773 | 0.118 |
| 12 | 113344953 | rs150799822 | A | G | 5.57E-06 | -1.4072 | 0.309801 | 0.1188 |
| 12 | 113345412 | rs111798104 | C | T | 1.13E-06 | -1.48579 | 0.305246 | 0.1203 |
| 12 | 113346021 | rs112606094 | G | A | 1.22E-06 | -1.48457 | 0.305932 | 0.1188 |
| 14 | 100278133 | rs541153352 | TA | T | 3.73E-06 | -1.47762 | 0.319436 | 0.1513 |
| 16 | 837731 | rs62031079 | T | C | 2.17E-06 | -1.4034 | 0.29624 | 0.1422 |
| 16 | 837945 | rs142659817 | T | C | 2.16E-06 | -1.40337 | 0.296231 | 0.1415 |
| 16 | 837953 | rs144270616 | C | A | 2.16E-06 | -1.40336 | 0.296231 | 0.1415 |
| 16 | 838512 | rs74631367 | A | T | 2.16E-06 | -1.40357 | 0.296266 | 0.1422 |
| 16 | 839087 | rs9652787 | C | T | 1.93E-06 | -1.40025 | 0.294161 | 0.1558 |
| 16 | 839248 | rs9972769 | C | G | 2.11E-06 | -1.40394 | 0.29601 | 0.1422 |
| 16 | 839253 | rs62012366 | T | C | 2.08E-06 | -1.39589 | 0.29418 | 0.1422 |
| 16 | 840155 | rs62012367 | A | G | 7.37E-07 | -1.37997 | 0.278712 | 0.1785 |
| 16 | 840260 | rs62012368 | T | C | 7.42E-07 | -1.38228 | 0.279245 | 0.1785 |
| 16 | 840308 | rs62012369 | G | C | 7.38E-07 | -1.37995 | 0.278712 | 0.1785 |
| 16 | 840327 | rs62012370 | A | G | 6.77E-07 | -1.37772 | 0.277325 | 0.1785 |
| 16 | 840525 | rs62012371 | T | C | 4.59E-06 | -1.54445 | 0.337002 | 0.1339 |
| 16 | 840643 | rs62012372 | A | G | 2.15E-06 | -1.39558 | 0.294479 | 0.1369 |
| 16 | 841006 | rs62012373 | T | C | 2.70E-06 | -1.38656 | 0.295503 | 0.1415 |
| 16 | 841050 | rs62012374 | G | A | 5.32E-06 | -1.32933 | 0.29204 | 0.1483 |

| | | | | | | | | |
|---|---|---|---|---|---|---|---|---|
| 16 | 844345 | rs62012403 | G | T | 9.24E-06 | -1.14149 | 0.257422 | 0.2118 |
| 16 | 20700594 | rs163254 | C | T | 5.96E-06 | 1.026428 | 0.226692 | 0.354 |
| 16 | 20706038 | rs121564 | T | G | 8.89E-06 | 0.996751 | 0.224359 | 0.3563 |
| 16 | 20728999 | rs4783487 | T | C | 9.02E-06 | -0.99817 | 0.224842 | 0.3533 |
| 16 | 31357495 | rs11574635 | C | G | 5.41E-06 | -1.23263 | 0.271006 | 0.2201 |
| 16 | 31359444 | rs13336235 | A | G | 5.48E-06 | -1.23185 | 0.271009 | 0.2239 |
| 19 | 30250716 | rs12972995 | C | G | 9.47E-06 | 1.150601 | 0.259787 | 0.1974 |
| 20 | 38649969 | rs220522 | A | T | 8.92E-06 | 1.479049 | 0.332988 | 0.118 |
| 20 | 38650180 | rs220521 | C | T | 6.14E-06 | 1.175569 | 0.25999 | 0.1604 |
| 20 | 38651713 | rs4812127 | G | T | 9.04E-06 | 1.477578 | 0.332864 | 0.1188 |
| 20 | 38666240 | rs220503 | T | C | 5.93E-06 | 1.41575 | 0.312595 | 0.1331 |
| 20 | 38669217 | rs220498 | T | C | 6.08E-06 | 1.41587 | 0.313008 | 0.1316 |
| 21 | 21626716 | rs1999296 | T | C | 7.29E-06 | -1.09354 | 0.243822 | 0.2844 |
| 22 | 50573113 | rs17001616 | A | C | 8.26E-06 | -1.3952 | 0.312943 | 0.2088 |

**Supplemental Table 2. Results from Selection Analysis**

| uniqID | Esan in Nigeria | Yoruba in Nigeria | Luhya in Kenya | Mende in Sierra Leone | Gambian Western Division | African Caribbean Barbados | African Ancestry Southwest |
|---|---|---|---|---|---|---|---|
| 12:113782624:A:G | x | x | | | | x | x |
| 12:113782758:A:G | x | x | | | | x | x |
| 12:113783217:C:T | x | x | x | x | | x | x |
| 12:113783826:A:G | x | x | x | x | | x | |

**Supplemental Table 3. Results from eQTL query**

| gene | snp_hg38 | pval |
|---|---|---|
| RRNAD1 | chr1:156792798:A:C | 0.022 |
| RRNAD1 | chr1:156788810:C:T | 0.03 |
| RRNAD1 | chr1:156736467:C:T | 0.036 |
| NARFL | chr16:837731:C:T | 0.045 |
| NARFL | chr16:837945:C:T | 0.045 |
| NARFL | chr16:837953:A:C | 0.045 |
| NARFL | chr16:839253:C:T | 0.045 |
| NARFL | chr16:840525:C:T | 0.045 |
| NARFL | chr16:841006:C:T | 0.045 |
| NARFL | chr16:841050:A:G | 0.045 |
| NARFL | chr16:840260:C:T | 0.048 |
| NARFL | chr16:840308:C:G | 0.048 |

**Commented [CS1]:** Ask Penny which column to use for build

**Commented [CS2R1]:** Use build 38.

**Supplemental Table 4 – TB candidate gene results**

| | Total SNPs | SNPs with P-value < 0.05 | %of total SNPs | Most Sig P-value |
|---|---|---|---|---|
| CD209 | 87 | 1 | 1.15 | 0.0217 |
| HLA-A | 278 | | | |
| HLA-DQB1 | 299 | | | |
| HLA-DRB1 | 145 | | | |
| IFNG | 22 | 1 | 4.55 | 0.0471 |
| IFNG-AS1 | 558 | 11 | 1.97 | 0.0048 |
| IFNGR1 | 18 | | | |
| IL10 | 23 | | | |
| MBL2 | 49 | 9 | 18.37 | 0.0038 |
| MCP1 | 20 | | | |
| SLC11A1 | 42 | 8 | 19.05 | 0.0055 |
| TLR2 | 52 | 3 | 5.77 | 0.0043 |
| TLR4 | 59 | 6 | 10.17 | 0.0189 |
| TLR9 | 13 | 2 | 15.38 | 0.0331 |
| TNFA | 19 | 1 | 5.26 | 0.0285 |
| VDR | 249 | 13 | 5.22 | 0.0070 |

**Supplemental Table 5 – Previously studied host-pathogen interaction effects in TB**

| Gene | Paper ref | Study population | Associated lineage | # SNPs/total SNPs p<0.05 | most sig p-val |
|---|---|---|---|---|---|
| MARCO | Thuong | Vietnam | Beijing | 0.0000 | 0.0780 |
| TLR2 | Caws | Vietnam | Beijing | 0.0857 | 0.0043 |
| FLOT1 | Luo | Peru | L2 | 0.0000 | 0.0608 |
| CD53 | Omae | Thailand | Beijing | 0.5479 | 0.00003 |
| HLA-B | Salie | South Africa | Beijing | 0.0000 | 0.1090 |
| HLA-B | Salie | South Africa | Beijing | 0.2736 | 0.0006 |
| HLA-B | Toyo-oka | Thailand | "Modern" | 0.0000 | 0.0588 |
| SLC11A1 | van Crevel | Indonesia | Beijing | 0.2800 | 0.0055 |
| IRGM | Intermann | Ghana | Mtb vs M africanum | 0.0000 | 0.0623 |
| LAMP1 | Songame | Indonesia | Beijing | 0.0000 | 0.2114 |
| MTOR | Songame | Indonesia | Beijing | 0.0000 | 0.0784 |
| DAP | Phelan | Thailand | L2.2 | 0.0000 | 0.0906 |
| DAP | Phelan | Thailand | L2.2 | 0.0000 | 0.1572 |
| chr14intergenic | Phelan | Thailand | L4.4 | 0.0556 | 0.0400 |
| MFAP2 | Phelan | Thailand | L2.2 | 0.0000 | 0.0774 |
| MFAP2 | Phelan | Thailand | L2.2 | 0.0000 | 0.2247 |
| FSTL5 | Phelan | Thailand | L2.2 | 0.0397 | 0.0036 |
| FSTL5 | Phelan | Thailand | L2.2 | 0.1000 | 0.0015 |
| chr2intergenic | Phelan | Thailand | L1.1 | 0.0000 | 0.1378 |
| RIMS3 | Phelan | Thailand | L1.1 | 0.0000 | 0.1148 |
| RIMS3 | Phelan | Thailand | L1.1 | 0.6000 | 0.0077 |
| chr3intergenic | Phelan | Thailand | L4.4 | 0.0000 | 0.1619 |
| CSGALNACT1 | Phelan | Thailand | L2.2 | 0.0127 | 0.0061 |
| CSGALNACT1 | Phelan | Thailand | L2.2 | 0.0000 | 0.1673 |

**Supplemental Figure 1. Correlation between GWAS findings for L4-Uganda vs L4-NonUganda (x-axis) and L4-Uganda vs all other lineages (y-axis), for results in primary GWAS where p < 0.005**

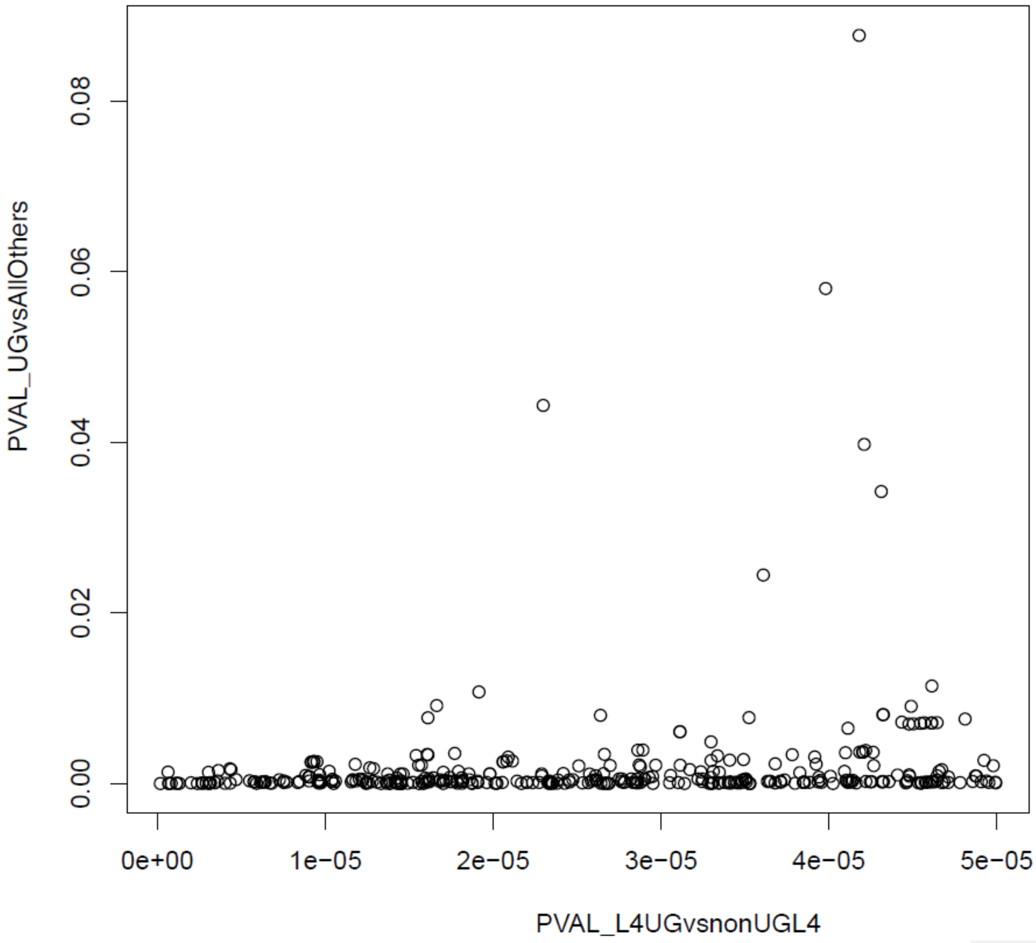
